# Development of iADJUST: a theory-informed, patient co-designed digital psychological intervention for adjustment in chronic kidney disease

**DOI:** 10.64898/2026.06.10.26355356

**Authors:** Pooja Schmill, Joanna L Hudson, Emily McBride, Sharlene Greenwood, Joseph Chilcot

## Abstract

**Background:** Psychological distress is common in chronic kidney disease (CKD) and associated with reduced quality of life, treatment non-adherence, and worse clinical outcomes. Distress in CKD is also linked to difficulties adjusting to the demands of illness management. Despite this, psychological support remains inconsistently integrated within kidney care pathways, and existing interventions often lack clear theoretical specification and explicit targeting of mechanisms underpinning adjustment to CKD.

**Objectives:** To describe the systematic development of iADJUST, a theory-informed patient co-designed digital psychological intervention targeting key cognitive and behavioural mechanisms involved in adjustment to CKD.

**Methods:** Intervention development was guided by the Medical Research Council framework for complex interventions. A structured, iterative process integrated empirical evidence, psychological theory, and patient and public involvement and engagement. The Common-Sense Model of Self-Regulation and cognitive behavioural theories informed the identification of modifiable maintaining mechanisms associated with adjustment to CKD. Intervention components were mapped onto these mechanisms and refined through co-design with people living with CKD.

**Results:** iADJUST is a six-session self-guided digital psychological intervention delivered over 12 weeks and supplemented by therapist contact. The intervention targets illness-related uncertainty, fatigue-related activity dysregulation, catastrophic “what-if” thinking, self-critical evaluation, and behavioural withdrawal. It integrates psychoeducation, cognitive and behavioural strategies, maintenance planning, and elements from acceptance and commitment therapy and compassion-focused approaches. Content is delivered through video, audio, and guided tasks and activities.

**Conclusion:** iADJUST provides a theory-informed, evidence-based psychological intervention for CKD explicitly mapping intervention components to maintaining cognitive and behavioural mechanisms implicated in adjustment. Feasibility evaluation is underway.

## Background

Chronic kidney disease (CKD) is associated with a substantial and persistent psychological burden, with high rates of depression and anxiety reported across disease stages [1, 2]. These challenges are clinically consequential, as psychological distress has been consistently associated with poorer quality of life, reduced engagement in self-management behaviours, and adverse health outcomes [3, 4]. In long-term conditions such as CKD, distress may also arise from difficulties adjusting to the emotional, cognitive, and behavioural demands of illness management [5, 6]. Despite this, psychological care within kidney services across the UK remains varied and fragmented, with limited access to structured, evidence-based interventions [3, 7, 8]. Digital approaches offer a potential means of addressing this gap by improving accessibility and scalability of psychological support within routine care.

This gap reflects a broader limitation within the field. While the importance of improving psychological outcomes in CKD is recognised as a clinical priority [7, 8], few interventions systematically target the cognitive and behavioural mechanisms through which CKD-specific distress and reduced functioning are maintained [9]. Explicit identification and targeting of these mechanisms are important for improving intervention precision, enhancing efficacy, and supporting evaluation of how and for whom interventions are effective.

Adjustment to CKD extends beyond the reduction of psychological symptoms alone. Individuals are required to manage ongoing uncertainty regarding disease progression, fluctuating symptoms such as fatigue, and changes in functional capacity [6, 10] Consequently, psychological intervention in CKD must address not only mood-related symptoms, but also broader processes underpinning adaptation to chronic illness [5, 8]. These include behavioural engagement, responses to symptom burden and treatment burden, and the maintenance of daily functioning, as these demands may give rise to interacting cognitive and behavioural processes that perpetuate distress and reduced quality of life [8]. Previous work has identified key processes, including negative illness representations, symptom burden (particularly fatigue), worry about future deterioration, behavioural withdrawal, and disrupted activity patterns, as central to adjustment trajectories in CKD [6, 11-13]. However, these processes have rarely been explicitly specified and linked to intervention components within CKD intervention development, limiting the ability to design targeted, testable, and scalable interventions.

A further limitation within the existing literature is the lack of interventions delivered earlier in the CKD pathway [9]. Much of the current evidence focuses on later-stage disease or dialysis populations [14], despite growing recognition of the importance of early preventative approaches [7, 8]. Intervening earlier in the disease trajectory has the potential to support psychological adjustment, maintain behavioural engagement, and promote self-management behaviours before patterns of distress become entrenched. Scalable digital interventions may be particularly well suited to early-stage delivery, offering opportunities to reach larger populations within routine care. Such an approach aligns with broader preventative and stepped-care models within the UK National Health Service (NHS) and may have implications for longer-term clinical outcomes.

This lack of theoretically grounded, mechanism-focused and early-stage interventions is reflected in the current evidence base [9, 14]. Our recent systematic review identified substantial heterogeneity in psycho-behavioural interventions for CKD, with inconsistent effects across psychological and behavioural outcomes and limited specification of active ingredients underpinning intervention effects [9]. In addition, few studies reported meaningful patient and public involvement and engagement (PPIE) in intervention development, limiting the relevance and acceptability of intervention content. Similarly, empirical data from kidney units in the NHS have demonstrated variability in the identification and management of psychological distress, with no standardised pathway for intervention [7, 8].

Collectively, these findings indicate the need for interventions that explicitly target modifiable cognitive and behavioural processes implicated in psychological adjustment to CKD, are delivered earlier in the disease trajectory, and are informed by patient perspectives. Digital delivery provides an opportunity to operationalise such approaches at scale. The present paper describes the development of iADJUST, a theory-informed patient-co-designed digital psychological intervention, to address this gap.

A key limitation of existing interventions in CKD is the limited specification of mechanisms underpinning psychological adjustment and their linkage to “active ingredients” included within the content of the intervention. iADJUST addresses this limitation by identifying cognitive and behavioural maintaining processes implicated in adjustment to CKD and transparently mapping these processes to intervention components with input from PPIE.

## Methods

### Intervention Development Approach

The development of iADJUST, a primarily self-guided digital psychological intervention, was informed by the Medical Research Council (MRC) framework for complex interventions [15, 16], which emphasises integrating theory, empirical evidence, and stakeholder input within an iterative development process. This approach supports the systematic identification of intervention targets and maintaining processes prior to evaluation.

The development process comprised four interrelated stages: (1) needs assessment; (2) specification of intervention targets and maintaining processes; (3) selection and operationalisation of intervention components; and (4) PPIE.

### 1. Needs assessment

The needs assessment was informed by three sources: empirical work conducted by the study team (MoodMaps study) [7], a systematic review of psycho-behavioural interventions in CKD [9], and PPIE.

Findings from the MoodMaps study [7] highlighted psychological distress, fatigue, and emotional well-being as key unmet needs within CKD care. These findings are supported by further evidence identifying fatigue as a major symptom burden in CKD [6, 10], which informed the inclusion of intervention components targeting fatigue-related activity dysregulation and mood-related behavioural withdrawal. The MoodMaps study also identified variability in access to psychological support across kidney services and highlighted the need for psychological intervention earlier in the CKD pathway, before patterns of distress become more established [7].

The systematic review identified substantial heterogeneity across psycho-behavioural interventions in CKD, with inconsistent effects on psychological outcomes and limited specification of intervention mechanisms [9]. Interventions rarely included intervention components that explicitly targeted activity regulation [9], despite behavioural engagement and fatigue being central challenges reported across CKD populations [6, 10]. In addition, much of the existing intervention literature focused on later-stage CKD or dialysis populations, highlighting the relative lack of early-stage psychological interventions [9, 14]. These findings informed the inclusion of pacing and behavioural activation components in iADJUST, alongside the positioning of the intervention earlier within the CKD pathway.

PPIE further reinforced the need for psychological support from the point of CKD diagnosis and provided additional insight into the lived experience of CKD. Contributors consistently described uncertainty regarding disease progression, fluctuating fatigue, behavioural withdrawal, reduced functioning, and self-critical responses to illness-related limitations. Several contributors described difficulties accessing psychological support until distress had significantly escalated. These findings informed the identification of candidate cognitive and behavioural maintaining processes relevant to psychological adjustment in CKD.

### 2. Specification of intervention target mechanisms

The specification of intervention targets was informed by psychological theories relevant to adjustment in long-term conditions, rather than formal behavioural analysis frameworks. Although behavioural analysis approaches are commonly recommended within intervention development frameworks such as the MRC guidance, these are typically applied when the primary target is health behaviour change [15, 16]. In contrast, iADJUST was developed primarily to address psychological distress and adjustment difficulties associated with CKD. Consequently, theoretical approaches relevant to emotional adjustment and self-regulation were prioritised to identify modifiable cognitive and behavioural processes underpinning psychological distress and adjustment difficulties.

The Common-Sense Model of Self-Regulation [17] was used to conceptualise illness representations and their influence on coping responses and emotional adjustment. Cognitive behavioural models, including social cognitive theory [18-20], informed the identification of maintaining processes, such as worry–avoidance cycles, behavioural withdrawal, fatigue-related activity dysregulation, self-critical thinking and low self-efficacy.

Based on this integration, the intervention was designed to target: (i) illness-related uncertainty and perceived control; (ii) fatigue-related activity dysregulation; (iii) catastrophic “what-if” thinking; (iv) self-critical evaluation; (v) behavioural withdrawal and reduced reinforcement; and (vi) self-regulation processes relevant to maintenance and relapse prevention.

### 3. Selection and operationalisation of intervention components

Intervention components were selected and operationalised to target the identified cognitive and behavioural maintaining processes underpinning psychological adjustment in CKD. Previous work in dialysis populations has demonstrated the utility of conceptualising these processes within a cognitive behavioural framework [11]. Accordingly, intervention content was grounded primarily in cognitive behavioural principles [19, 20], with selected techniques from acceptance and commitment therapy [21, 22] and compassion-focused approaches [23] incorporated where theoretically relevant.

Psychoeducation was used to support illness understanding and normalise emotional responses. Behavioural strategies, including pacing and behavioural activation, targeted fatigue-related activity dysregulation, behavioural withdrawal and low perceived self-efficacy. Cognitive techniques were incorporated to address catastrophic ‘what-if’ thinking and illness-related uncertainty [19, 20]. Selected strategies informed by acceptance and commitment therapy [21, 22] and compassion-focused therapy [23] were also integrated to address psychological flexibility against a backdrop of illness-related uncertainty and self-critical evaluation. These approaches have previously been applied within long-term condition (LTC) populations, including individuals living with CKD [24, 25]. Maintenance planning was also incorporated to support longer-term self-regulation and consolidation of skills. All components were integrated within a cognitive behavioural framework rather than delivered as stand-alone therapeutic approaches.

### 4. PPIE

PPIE was embedded throughout the development process to ensure a person-centred approach and enhance the acceptability and relevance of the digital intervention. [26, 27]. Eight individuals with lived experience of CKD were recruited through kidney charities and social media platforms, including Instagram, Facebook, and LinkedIn.

As seen in Table 1, PPIE contributors represented a range of demographic and clinical experiences across the CKD pathway, including dialysis and transplant experience as well as varying levels of involvement in research or advisory activities. Several contributors also reported prior experiences of psychological distress and difficulties accessing psychological support related to CKD.

**Table 1.** Characteristics of PPIE contributors.

| **Characteristic** | **n (%)** |
| --- | --- |
| **Total PPIE contributors** | 8 |
| **Age range** |  |
| 25–34 years | 3 (37.5%) |
| 45–54 years | 3 (37.5%) |
| ≥55 years | 2 (25.0%) |
| **Sex** |  |
| Male | 6 (75.0%) |
| Female | 2 (25.0%) |
| **Ethnicity** |  |
| White British/Irish/Other | 5 (62.5%) |
| Black British | 2 (25.0%) |
| Asian British | 1 (12.5%) |
| **CKD stage/experience** |  |
| Early-stage CKD (Stages 1–3) | 2 (25.0%) |
| Later-stage CKD (Stages 4–5, non-dialysis) | 3 (37.5%) |
| Dialysis | 1 (12.5%) |
| Kidney transplant | 2 (25.0%) |
| **Previous psychological support for CKD-related challenges** |  |
| Yes | 5 (62.5%) |
| No | 3 (37.5%) |
| **Reported challenges accessing psychological support** |  |
| Yes | 3 (37.5%) |
| No | 5 (62.5%) |
| **Previous involvement in advisory/workshop activities** |  |
| First-time contributors | 3 (37.5%) |
| Occasional involvement | 3 (37.5%) |
| Frequent involvement | 2 (25.0%) |

An initial two-hour online workshop via video conferencing explored the lived experiences of CKD and identified key psychological and practical challenges. Participants were asked open-ended questions relating to emotional impact, support needs, and preferences for digitally delivered intervention support. The workshop was structured to ensure all participants could contribute, with facilitated discussion and opportunities for both verbal and written input. Findings from this workshop informed the initial structure and content of the intervention.

A draft intervention framework was subsequently developed and shared with PPIE contributors through structured questionnaires to gather feedback on relevance, acceptability, feasibility, and perceived burden. This feedback informed refinement of session structure, format (e.g., video, audio, activities and tasks), and overall design for a digital, self-guided intervention.

Full session scripts were then developed and reviewed iteratively. Each PPIE contributor provided detailed feedback on assigned sessions, focusing on clarity, tone, relevance, and manageability. Feedback was used to refine content before finalisation. This iterative process supported the incorporation of diverse patient perspectives and ensured that the intervention was acceptable, feasible, aligned with patient priorities, and optimised for digital delivery.

### Intervention format and delivery

The intervention comprises six approximately 30-minute digital sessions delivered in a primarily self-guided format over 12 weeks. Sessions are supplemented by six brief fortnightly contacts with a therapist to support engagement with the intervention. Content is delivered through a combination of video-based psychoeducation, audio-guided experiential tasks, and structured behavioural activities to support skill acquisition and real-world application.

A digital format was selected to maximise accessibility and scalability, while the intervention dosage aligns with low-intensity cognitive behavioural approaches within NHS services [28]. The use of multiple digital formats (video, audio, and interactive activities and tasks) was informed by PPIE feedback to enhance engagement, usability, and acceptability.

### Integration within CKD care pathways

iADJUST was developed as a psychological intervention intended to integrate within existing CKD care pathways, including the Kidney BEAM programme, a digital physical activity-based intervention that has demonstrated improvements in physical functioning and mental health-related quality of life [29].

iADJUST is conceptualised as a complementary intervention targeting cognitive and behavioural processes relevant to psychological adjustment and self-management in CKD. By addressing factors such as fatigue-related activity dysregulation, low mood, and illness-related uncertainty, the intervention is intended to support engagement in health-promoting behaviours, including physical activity, within routine care.

## Results

### Overview of the iADJUST intervention

iADJUST was designed as a primarily self-guided digital psychological intervention supported by brief therapist contact. The intervention aligns with low-intensity cognitive behavioural approaches used within NHS Talking Therapies services [28], balancing therapeutic depth with feasibility, scalability, and accessibility within routine care.

Each session targets a specific cognitive or behavioural maintaining process implicated in adjustment to CKD, with content sequenced to support progressive development of illness understanding, behavioural regulation, and longer-term self-regulation. The mapping of maintaining processes to intervention components, therapeutic approaches, and delivery formats is summarised in Table 2.

**Table 2.**
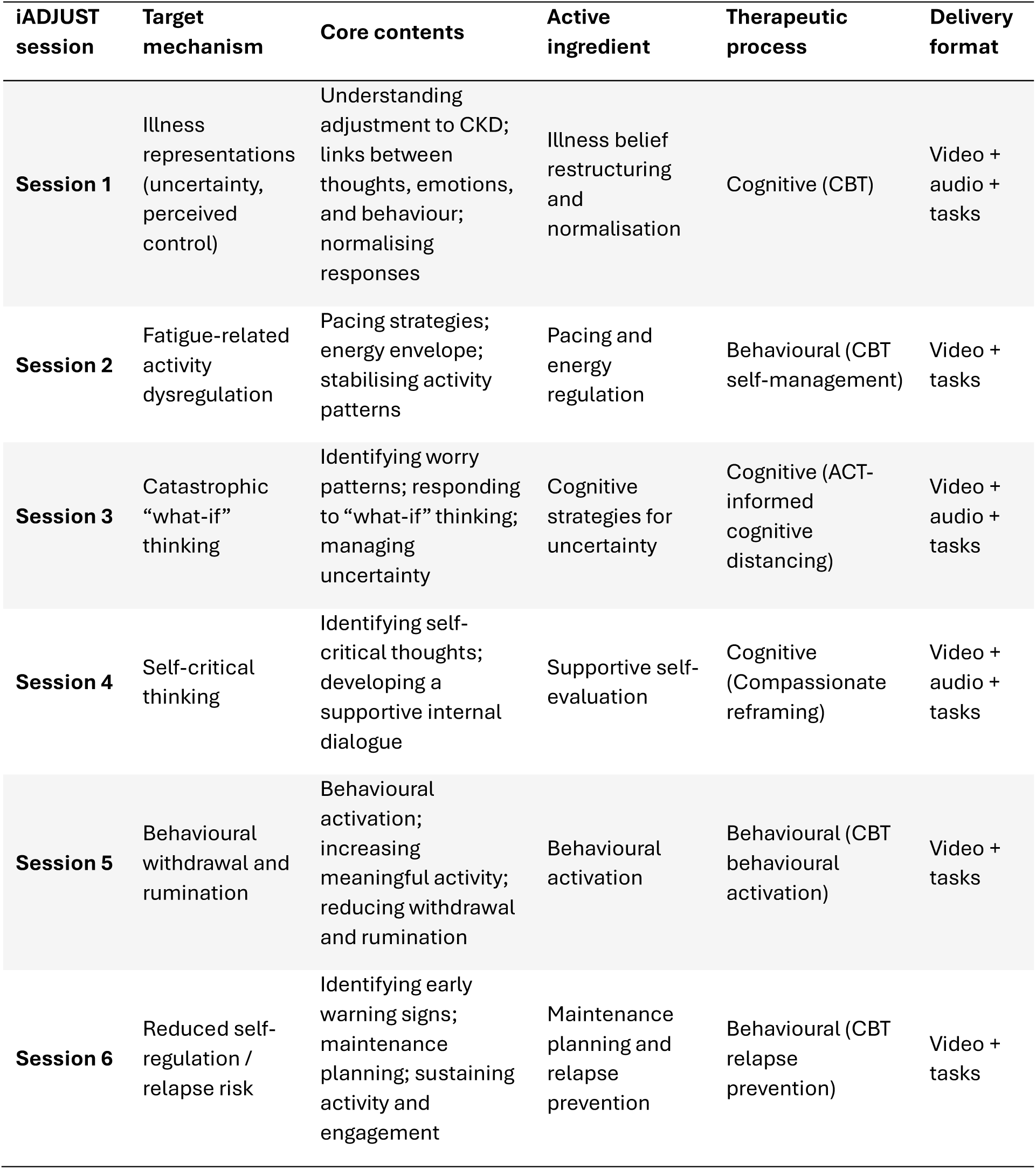
Mapping of target mechanisms to intervention functions and active ingredients.

### Intervention delivery and structure

Across sessions, content is delivered within a self-guided digital format through structured psychoeducation (video-based), guided experiential tasks (audio-based), and applied behavioural activities designed to support skill acquisition and real-world implementation. Intervention materials are interactive and practice-oriented, enabling participants to apply strategies between sessions and integrate skills into everyday self-management.

Sessions were sequenced to support progression from understanding of the illness and behavioural stabilisation to cognitive and behavioural skill development, followed by maintenance and relapse prevention.

A logic model was developed to represent the hypothesised relationships between CKD-related challenges, maintaining cognitive and behavioural processes, intervention components, and outcomes (Fig 1). The model illustrates how intervention components are intended to influence modifiable cognitive and behavioural maintaining processes associated with psychological adjustment, including unhelpful common-sense illness beliefs about CKD, illness-related uncertainty, behavioural withdrawal, fatigue-related activity dysregulation, and self-critical thinking. These proximal changes are hypothesised to support improvements in psychological well-being, behavioural engagement, and participation in health-promoting activities.

**Fig 1.**
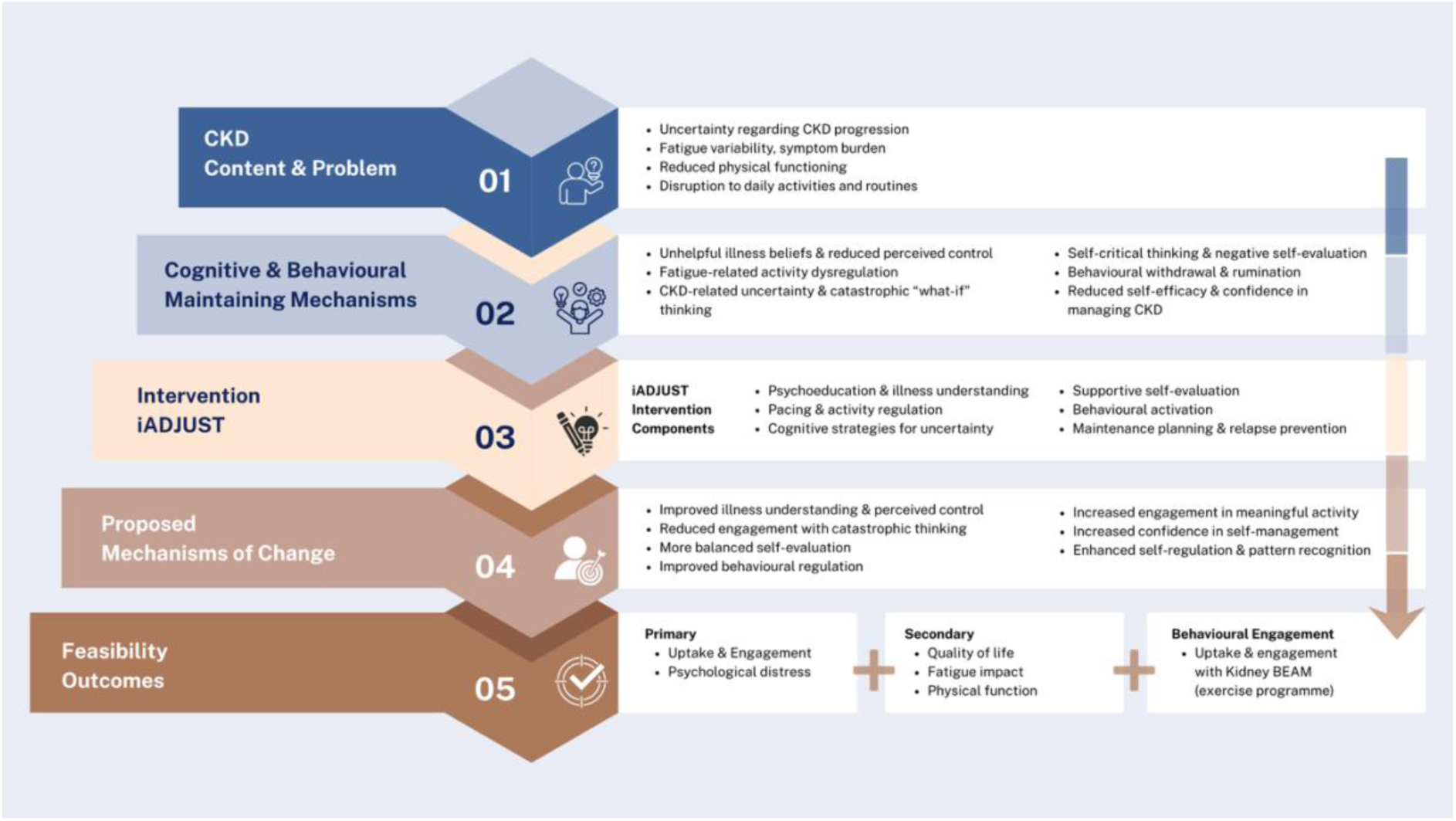
Logic model of the iADJUST intervention.

The logic model, therefore, provides a testable framework linking intervention components to both intermediate mechanisms of change and downstream behavioural and clinical outcomes.

## Discussion

This paper describes the development of a theory-informed, mechanism-focused digital psychological intervention targeting adjustment to CKD. A central contribution of this work is the explicit specification of cognitive and behavioural mechanisms underpinning psychological distress and their direct linkage to intervention components within a coherent theoretical framework.

Existing psycho-behavioural interventions in CKD have been characterised by heterogeneity in content and limited specification of mechanisms underpinning intervention effects [9]. In contrast, iADJUST was developed through the integration of the Common-Sense Model [17] and cognitive behavioural theories [18-20] to identify and target specific maintaining mechanisms, including illness-related uncertainty, catastrophic “what-if” thinking, self-critical evaluation, behavioural withdrawal, low self-efficacy and fatigue-related activity dysregulation. This approach builds on previous model-driven intervention development work conducted in end-stage CKD [11]. The current intervention extends these principles to earlier stages of the CKD pathway, incorporating behavioural regulation alongside illness representations and cognitive processes. By explicitly linking intervention techniques to modifiable maintaining mechanisms, this approach has the potential to improve both the precision and efficiency of intervention design, and to support the identification of active ingredients responsible for change.

Importantly, specification of mechanisms of action enables the generation of testable hypotheses regarding pathways of change. This moves beyond descriptive accounts of intervention development towards a more explanatory, mechanism-driven approach, which is essential for advancing intervention science in CKD. It also enables examination of for whom interventions are most effective, which is particularly important in CKD populations characterised by clinical and demographic heterogeneity [2, 8].

A further contribution lies in the positioning of iADJUST within an integrated model of CKD care. Rather than conceptualising psychological support as an adjunct, the intervention is embedded early in the CKD pathway and targets processes which are likely to influence engagement with core self-management behaviours such as physical activity. This represents an important shift from existing CKD interventions, which have largely focused on end-stage CKD or dialysis populations [14], towards a more preventative approach that targets adjustment processes before unhelpful cognitive and behavioural patterns become entrenched.

By addressing cognitive and behavioural barriers such as fatigue-related dysregulation, low mood, and illness-related uncertainty, iADJUST may enhance the effectiveness of downstream interventions and support more comprehensive person-centred models of care. Early intervention at scale, particularly through digital delivery, has the potential to improve longer-term psychological and behavioural outcomes, although this requires evaluation in future longitudinal studies.

The intervention also reflects developments in psychological therapy approaches. In addition to core cognitive behavioural principles, iADJUST incorporates elements from third-wave cognitive behavioural therapies, including acceptance and commitment therapy [21, 22] and compassion-focused approaches [23]. These approaches target cognitive processes such as cognitive fusion, experiential avoidance, and self-critical thinking, which are increasingly recognised as important in the maintenance of distress across long-term conditions [21-23, 30]. Their inclusion reflects advances in the field and extends beyond traditional CBT approaches used in earlier CKD interventions [11].

The integration of PPIE throughout the development process ensured that the intervention was grounded in lived experience and responsive to patient priorities. This is particularly important in the context of self-guided, digital interventions where acceptability, relevance, and usability are critical determinants of engagement and adherence [26]. The inclusion of patient perspectives throughout development also addresses a recognised limitation in previous CKD interventions, where patient input has often been limited or absent [9].

The accompanying logic model provides a structured representation of the hypothesised relationships between CKD-related challenges, cognitive and behavioural maintaining mechanisms, intervention components, and outcomes. This facilitates both the evaluation of effectiveness and the examination of mechanisms of change, which remains a key gap in the current literature.

Taken together, this work advances the development of psychological interventions in CKD by demonstrating how psychological theory, empirical evidence, and input from people with lived experience can be systematically integrated to produce a coherent, testable, and clinically relevant intervention. By embedding explicit specifications of maintaining mechanisms and contemporary therapeutic approaches within intervention design using digital formats, iADJUST provides a framework for developing scalable psychological interventions that can be implemented earlier in the disease pathway and evaluated using theory-driven methods.

## Conclusion

iADJUST is a theory-informed, evidence-based, and patient co-designed digital psychological intervention targeting key cognitive and behavioural mechanisms underpinning adjustment to CKD. By explicitly linking maintaining cognitive and behavioural mechanisms to intervention components, the programme provides a transparent and testable framework for understanding how psychological support may influence both emotional well-being and engagement in self-management behaviours. As such, iADJUST addresses a critical gap in kidney care, where psychological needs are increasingly recognised but remain inconsistently addressed through structured, mechanism-focused interventions.

Designed as a primarily self-guided digital intervention, supported by brief therapist contact, iADJUST offers a potentially scalable approach to integrating psychological support earlier within CKD care pathways. Ongoing feasibility evaluation will assess the acceptability, engagement, and implementation potential of the intervention, informing its refinement and future testing within routine kidney care. If feasible and acceptable, iADJUST may provide a foundation for larger-scale evaluation of early, theory-driven psychological intervention in CKD.

## Acknowledgements

This study is funded by the National Institute for Health and Care Research (NIHR) Maudsley Biomedical Research Centre (BRC). The views expressed are those of the author(s) and not necessarily those of the NHS, the NIHR or the Department of Health and Social Care. The authors gratefully express their appreciation to all study authors who responded to our request for additional data.

## Declarations

### Ethical approval

This study describes the development of a digital psychological intervention co-designed by patient and public involvement and engagement (PPIE). PPIE activities were conducted to inform intervention design and refinement, and did not constitute research procedures requiring formal ethical approval.

### Informed consent to participate

All PPIE contributors provided informed consent to participate in intervention development activities and for their feedback to be used to inform the design and refinement of the intervention.

### Data availability statement

No datasets were generated or analysed during the current study. Materials relating to the intervention development process are available from the corresponding author upon reasonable request.

### Funding acknowledgement

This work was supported by the National Institute for Health and Care Research (NIHR) Maudsley Biomedical Research Centre (BRC). The views expressed are those of the authors and not necessarily those of the NHS, the NIHR, or the Department of Health and Social Care.

### Author contributions

P.S. conceived the study and led the intervention development, PPIE activities, data interpretation, and manuscript preparation. J.H. and E.M. contributed to intervention development, theoretical refinement, and critical revision of the manuscript. All authors reviewed and approved the final manuscript.

### Acknowledgements

The authors would like to thank the PPIE contributors who generously shared their time, experiences, and feedback throughout the development of iADJUST. Their contributions were instrumental in ensuring the intervention was relevant, acceptable, and responsive to the needs of people living with chronic kidney disease.

